# Have there been sustained impacts of the Covid-19 pandemic on trends in smoking prevalence, uptake, quitting, use of treatment, and relapse? A monthly population study in England, 2017-2022

**DOI:** 10.1101/2022.12.06.22283023

**Authors:** Sarah E. Jackson, Harry Tattan-Birch, Lion Shahab, Emma Beard, Jamie Brown

## Abstract

**Background:** This study aimed to examine whether there have been sustained impacts of the Covid-19 pandemic on smoking patterns in England.

**Methods:** Data were from 101,960 adults (≥18y) participating in a monthly representative household survey between June-2017 and August-2022. Interview were conducted face-to-face until March 2020 and via telephone thereafter. Generalised additive models estimated associations of the pandemic onset (March-2020) with current smoking, uptake, cessation, quit attempts, medium-term abstinence, and use of support. Models adjusted for seasonality, sociodemographic characteristics, and (where relevant) dependence and tobacco control mass-media expenditure.

**Findings:** Before the Covid-19 pandemic, smoking prevalence fell by 5.2% per year; this rate of decline slowed to 0.3% per year during the pandemic (RR_Δtrend_=1.06, 95%CI=1.02-1.09). This slowing was evident in more but not less advantaged social grades (RR_Δtrend_=1.15, 1.08-1.21; RR_Δtrend_=1.00, 0.96-1.05). There were sustained step-level changes in different age groups: a 34.9% (95%CI=17.7-54.7%) increase in smoking prevalence among 18-24-year-olds, indicating a potential rise in uptake, in contrast to a 13.6% (95%CI=4.4-21.9%) decrease among 45-65-year-olds. There were sustained increases in quitting among past-year smokers, with a 120.4% (95%CI=79.4-170.9%) step-level increase in cessation and a 41.7% (95%CI=29.7-54.7%) increase in quit attempts.

**Interpretation:** In England, the rate of decline in adult smoking prevalence stagnated during the Covid-19 pandemic. Potential reductions in smoking prevalence among middle-aged adults and sustained increases in quitting among smokers may have been offset by a sustained rise in uptake among young adults. The slowing in the rate of decline was pronounced in more advantaged social grades.

**Funding:** Cancer Research UK.

**Putting research into context:** *Evidence before this study:* We searched PubMed up to 1 September 2022 for papers on the Covid-19 pandemic and changes in smoking patterns among adults in England published since March 2020, using the terms “smok*” AND (“Covid*” OR “pandemic”) AND (“England” OR “UK”) AND “adults”. Of the 167 studies that were identified, none reported on trends in smoking behaviour among adults in England during the Covid-19 pandemic using more than two data points. Three prospective studies compared smoking and quitting behaviour before and during the initial stages of the pandemic using a pre-post design: two observed no notable change in smoking prevalence but documented increases in quit attempts and cessation during the first lockdown in England (April-July 2020); the other reported an uncertain decline in smoking prevalence in April 2020. Two prospective studies compared self-reported changes in consumption and attempts to quit at two time points, 12 months apart, during the pandemic; neither observed any change over time. Two cross-sectional studies and two qualitative studies described self-reported changes in smoking behaviour during the pandemic. The majority of other search results reported on the association between smoking status and risk of Covid-19 infection and outcomes; or included smoking status as a covariate in analyses of associations between other factors and the pandemic.

*Added value of this study:* This study uses a nationally representative survey of adults in the general population in England, conducted monthly over a 5-year period, to estimate sustained impacts of the Covid-19 pandemic on smoking patterns. Unlike previous studies, which have focused on changes that occurred early in the pandemic, this study includes data up to August 2022 (two and a half years after the pandemic started) which allows us to understand whether any initial changes in smoking and quitting behaviour have been sustained over time. By collecting data monthly, this study is the first of its kind to have a large enough number of data points to undertake this kind of analysis (most representative surveys collect data annually). The findings are of direct relevance to the UK government’s aim to reduce smoking prevalence in England to less than 5% by 2030 and should inform a new tobacco control plan in England.

*Implications of all the available evidence:* Before the Covid-19 pandemic, smoking prevalence had been falling among adults in England at a near linear rate for more than 20 years. This historic steady decline has almost completely stopped since the start of the pandemic. This may have been caused by a potential rise in uptake of smoking among young adults. These changes demand urgent, bold policy action, including measures to deter youth uptake of smoking and to support smokers to quit.

## Introduction

The Covid-19 pandemic had a profound impact on everyday life, public health, and health services. Studies conducted during the early stages of the pandemic documented mixed changes in smoking behaviour. Many observed short-term increases in rates of quit attempts and cessation among smokers,^1–4^ indicating the pandemic may have prompted smokers to stop. However, surveys that measured smoking prevalence produced inconsistent findings (both between and within countries), including increases, decreases, and no substantial change in the proportion of adults who smoke.^1,2,5^ Evidence on changes in smoking prevalence has been limited by many studies using non-representative samples^2^ and nationally representative surveys undergoing substantial changes to their methods of data collection and sample weighting as a result of social distancing restrictions.^5^ Relatively little is known about what impact the pandemic has had on uptake of smoking,^2,6^ relapse among ex-smokers, or use of support by smokers trying to quit^1^ (see Supplementary Material for a more detailed literature review).

Identifying whether any short-term changes in smoking patterns following the onset of the pandemic have translated into long-term, sustained changes, and the groups in which they have occurred, is essential for building a clear picture of its public health impact and targeting policy, messaging, and support services. As highlighted by Sarich et al.,^2^ there is some evidence that lifestyle behaviours adopted during a pandemic can persist for some time – for example, sustained increases in alcohol abuse/dependence symptoms were observed three years after the 2003 SARS outbreak among individuals in China who were quarantined or worked in high-risk settings during the epidemic.^7^ On the other hand, it is possible that once life started to return to ‘normal’ after the early months of the pandemic, people reverted to their previous smoking patterns and quitting became a less salient issue.

Two and a half years on from the start of the pandemic, there are now sufficient data within the Smoking Toolkit Study (a representative monthly survey of adults in England) to undertake a more detailed analysis of whether there has been a sustained impact of the Covid-19 pandemic on smoking patterns. In collecting data monthly, the Smoking Toolkit Study affords a unique opportunity to assess detailed trends at this stage (most representative surveys collect these data at much less frequent intervals). Specifically, we aimed to address the following research questions. Using data from June 2017 through August 2022:

1. What has been the sustained impact of the Covid-19 pandemic on monthly trends in:
  a. Current smoking (among all adults);
  b. Current smoking among young adults (to assess uptake of smoking);
  c. Current smoking among middle-aged adults (to gauge late relapse);
  d. Cessation and making ≥1 serious quit attempt (among past-year smokers);
  e. Number of past-year quit attempts and use of cessation support in the most recent attempt (among past-year smokers who made ≥1 quit attempt);
  f. Medium-term abstinence (indexed by success of quit attempts that started 6-12 months before the survey, to gauge medium-term relapse)?

We also explored the impact of the Covid-19 pandemic on these outcomes separately by socioeconomic position.

## Method

### Pre-registration

The analysis plan was pre-registered on Open Science Framework (https://osf.io/vy254/). We made one amendment: the model assessing medium-term relapse (i.e., failure of quit attempts that started 6-12 months prior to the survey) had problems with convergence due to a very high rate of relapse, so we inverted the coding for this outcome (1=0, 0=1) and reported results as medium-term abstinence (i.e., success of quit attempts that started 6-12 month prior).

### Design

The Smoking Toolkit Study uses a hybrid of random probability and simple quota sampling to select a new sample of 1,700 adults in England each month.^8^ Comparisons with sales data and other national surveys indicate that key variables including sociodemographics, smoking prevalence, and cigarette consumption are nationally representative.^8,9^

Data were collected monthly, initially through face-to-face computer-assisted interviews. However, social distancing restrictions under the Covid-19 pandemic meant no data were collected in March 2020 and data from April 2020 onwards were collected via telephone. The telephone-based data collection used the same combination of random location and quota sampling, and weighting approach as the face-to-face interviews and comparisons of the two data collection modalities indicate good comparability.^1,10,11^ Nonetheless, it will not be possible to determine with certainty whether any step-level changes observed are due to the pandemic or the switch from face-to-face to telephone interviewing. While step-level changes may have been affected, changes in the *slope* of trends from before to after the pandemic are likely unaffected — given that there were no further updates in methodology after April 2020.

For the present study, we used individual-level data collected between June 2017 and August 2022. Because the sample was restricted to people aged ≥18⍰years when data collection switched from face-to-face to telephone interviews, we excluded any participants aged 16–17 recruited before April 2020 for consistency.

### Measures

Full details of the measures (including question wording and derivation) are provided in Supplementary Material.

We assessed the following outcomes:

- Among all adults: ***current smoking***;
- Among 18-24 year-olds: current smoking (as an indicator of ***uptake***, because any increases in this age group would largely be driven by uptake rather than relapse);
- Among 45-65 year-olds: current smoking (as an indicator of ***late relapse***, because an increase in this age group would largely be driven by relapse rather than uptake.^12^ The upper age limit for this group was selected to minimise any impact of increased mortality at older ages during the pandemic on smoking prevalence);
- Among past-year smokers: ***cessation, making ≥1 serious quit attempt*** in the past year;
- Among past-year smokers who made ≥1 quit attempt in the past year: ***number of quit attempts*** made (log-transformed), ***use of prescription medication*** (varenicline/bupropion/nicotine replacement therapy), ***use of behavioural support*** (face-to-face support/telephone support/websites/apps/written self-help materials), ***use of e-cigarettes***;
- Among past-year smokers who made ≥1 quit attempt 6-12 months before completing the survey: ***medium-term abstinence***.

Covariates included age, gender, occupational social grade (ABC1=managerial/professional/intermediate, C2DE=small employers/lower supervisory/technical/semi-routine/routine/never workers/long-term unemployed), region in England, and (where relevant) level of dependence and government spending on tobacco control mass media campaigns.

### Statistical analysis

Data were analysed in R v.4.2.1. Missing cases were excluded on a per-analysis basis. We calculated unweighted and weighted descriptive statistics on sociodemographic and smoking characteristics. The Smoking Toolkit Study uses raking to match the sample to the population in England on the dimensions of age, social grade, region, housing tenure, ethnicity, and working status within sex.^8^ All the following analyses were done on weighted data.

We used segmented regression to assess the effect of the onset of the Covid-19 pandemic on each outcome. We used generalised additive models (GAMs). These allow the fitting of smoothing terms (e.g., cyclic cubic splines) to take seasonality into account. We modelled the trend in each outcome before the pandemic (underlying secular trend; coded 1…*n*, where *n* was the total number of waves), the step-level change (coded 0 before the start of the pandemic in March 2020 and 1 after), and change in the trend (slope) post-onset of the pandemic relative to pre-pandemic (coded 0 before the pandemic and 1…*m* from April 2020 onwards, where *m* was the number of waves after the start of the pandemic). Models were adjusted for seasonality (modelled using a smoothing term with cyclic cubic splines specified) and covariates. A linear pre-pandemic and pandemic trend was assumed, based on prior data^13^ and the relatively short length of the time-series (meaning we expected negligible differences between log-linear and linear trends). We repeated models separately by social grade (ABC1/C2DE). We used predicted estimates from these models to plot time trends in the weighted prevalence (or mean, for number of quit attempts) of each outcome alongside unadjusted, weighted monthly data points.

Planned sensitivity analyses tested for pulse effects by running GAMs with pulses lasting two and three months (coded 0 before the start of the pandemic, 1 in the two or three months after the onset of the pandemic, and 0 thereafter), assuming a constant underlying time trend. Next, we reran models for cessation, medium-term abstinence, and use of support excluding our measure of cigarette dependence (strength of urges to smoke) as a covariate, because this could plausibly be affected by the Covid-19 pandemic (e.g., increased due to stress or reduced due to less exposure to others smoking) and thus adjusting for it may have served to dilute the true impact of the pandemic on these outcomes. We also reran the model for use of prescription medication excluding varenicline, to check whether the results were affected by unavailability of this medication from mid-2021 due to manufacturer recall.

Finally, we included an unplanned analysis in which we modelled changes in cigarette dependence in relation to the Covid-19 pandemic (using GAMs as described above, with adjustment for age, gender, social grade, and region), to provide context on differences between analyses that did and did not include dependence as a covariate.

## Results

There were 102,371 respondents to the Smoking Toolkit Study between June 2017 and August 2022. We excluded 411 people who did not report their smoking status, leaving a sample of 101,960 for analysis. Of these, 55,349 were surveyed before the start of the pandemic (June 2017–February 2020) and 46,611 were surveyed during the pandemic (April 2020–August 2022). Table 1 presents weighted descriptive statistics for the sample as a whole and as a function of the timing of the pandemic (unweighted characteristics are shown in Supplementary Table 1).

**Table 1.** Descriptive statistics

|  | Overall | Before pandemic<br>(June 2017-Feb 2020) | During pandemic<br>(April 2020-Aug 2022) |
| --- | --- | --- | --- |
| *Unweighted N* | *101 960* | *55 349* | *46 611* |
| Age (years) | 48.3 (18.7) | 47.8 (18.9) | 48.7 (18.6) |
| 18-24 | 12.1% | 12.8% | 11.3% |
| 25-44 | 33.2% | 33.1% | 33.3% |
| 45-65 | 33.2% | 33.0% | 33.5% |
| >65 | 21.5% | 21.1% | 21.9% |
| Gender |  |  |  |
| Men | 48.9% | 49.0% | 48.7% |
| Women | 50.8% | 50.9% | 50.7% |
| In another way | 0.3% | 0.1% | 0.6% |
| Social grade C2DE | 44.2% | 44.4% | 43.9% |
| Region in England |  |  |  |
| London | 15.5% | 15.5% | 15.5% |
| South | 26.5% | 26.4% | 26.5% |
| Central | 30.2% | 30.2% | 30.3% |
| North | 27.8% | 27.9% | 27.7% |
| Current smoker |  |  |  |
| All adults | 16.8% | 17.0% | 16.6% |
| Young adults<sup>1</sup> | 22.7% | 21.7% | 24.0% |
| Middle-aged adults<sup>2</sup> | 15.2% | 16.5% | 13.5% |
| Past-year smoker | 18.5% | 18.1% | 19.0% |
| Cigarette dependence<sup>3a</sup> | 1.69 (1.19) | 1.74 (1.13) | 1.62 (1.26) |
| Quit in past year<sup>3</sup> | 9.2% | 6.2% | 12.6% |
| Tried to quit in past year<sup>3</sup> | 33.9% | 30.7% | 37.5% |
| Number of past-year quit attempts<sup>4b</sup> | 1.43 (1.79) | 1.40 (1.77) | 1.45 (1.79) |
| Medium-term abstinence<sup>5</sup> | 17.0% | 12.1% | 21.3% |
| Use of cessation support<sup>4</sup> |  |  |  |
| Prescription medication | 7.1% | 6.8% | 7.4% |
| Behavioural support | 7.4% | 6.1% | 8.7% |
| E-cigarettes | 31.3% | 32.7% | 29.8% |
| Monthly inflation-adjusted national tobacco control expenditure (£)<sup>c</sup> | 137 000<br>(247 000) | 142 000<br>(237 000) | 131 000<br>(261 000) |

Data are shown as percentages or mean (SD), unless otherwise specified.
<sup>1</sup> Among 18-24 year olds (*unweighted n* = 12,455). <sup>2</sup> Among 45-65 year olds (*unweighted n* = 34,332). <sup>3</sup> Among past-year smokers (*unweighted n* = 17,964). <sup>4</sup> Among past-year smokers who tried to quit in the past year (*unweighted n* = 5,754). <sup>5</sup> Among past-year smokers who made a quit attempt that started 6-12 months before completing the survey (*unweighted n* = 2,855).
<sup>a</sup> Strength of urges to smoke rated on a scale from 0 (none) to 5 (extremely strong).
<sup>b</sup> Geometric mean.
<sup>c</sup> Population-level variable, unweighted.
There was a small amount of missing data for some variables (<0.1% gender, 0.1% medium-term abstinence, 2.4% cigarette dependence, 4.1% tried to quit in past year); valid percentages are shown.

### Current smoking

Table 2 summarises the GAM results. Figure 1 shows trends in current smoking over the study period.

**Table 2.** GAM results: associations between the Covid-19 pandemic and smoking outcomes, overall and by social grade

|  | Overall |  |  | Social grades ABC1 |  |  | Social grades C2DE |  |  |
| --- | --- | --- | --- | --- | --- | --- | --- | --- | --- |
|  | RR / | 95% CI |  | RR / | 95% CI |  | RR / | 95% CI |  |
|  | *Exp. B* * | Lower | Upper | *Exp. B* * | Lower | Upper | *Exp. B* * | Lower | Upper |
| Current smoking<sup>1a</sup> |  |  |  |  |  |  |  |  |  |
| Trend (year) | 0.948 | 0.927 | 0.970 | 0.905 | 0.873 | 0.939 | 0.969 | 0.941 | 0.999 |
| Level | 1.080 | 1.022 | 1.141 | 1.201 | 1.101 | 1.310 | 1.027 | 0.953 | 1.106 |
| ΔTrend (year) | 1.052 | 1.014 | 1.090 | 1.145 | 1.083 | 1.211 | 1.003 | 0.955 | 1.054 |
| Current smoking among young adults<sup>2a</sup> |  |  |  |  |  |  |  |  |  |
| Trend (year) | 0.895 | 0.846 | 0.946 | 0.876 | 0.809 | 0.948 | 0.909 | 0.838 | 0.986 |
| Level | 1.349 | 1.177 | 1.547 | 1.369 | 1.127 | 1.663 | 1.342 | 1.101 | 1.636 |
| ΔTrend (year) | 1.072 | 0.982 | 1.171 | 1.138 | 1.004 | 1.290 | 1.018 | 0.896 | 1.158 |
| Current smoking among middle-aged adults<sup>3a</sup> |  |  |  |  |  |  |  |  |  |
| Trend (year) | 0.943 | 0.907 | 0.981 | 0.883 | 0.828 | 0.942 | 0.974 | 0.924 | 1.026 |
| Level | 0.864 | 0.781 | 0.956 | 1.069 | 0.909 | 1.257 | 0.776 | 0.674 | 0.893 |
| ΔTrend (year) | 1.096 | 1.025 | 1.172 | 1.171 | 1.055 | 1.300 | 1.058 | 0.964 | 1.162 |
| Cessation<sup>4b</sup> |  |  |  |  |  |  |  |  |  |
| Trend (year) | 0.839 | 0.761 | 0.926 | 0.926 | 0.820 | 1.047 | 0.755 | 0.645 | 0.884 |
| Level | 2.204 | 1.794 | 2.709 | 1.770 | 1.371 | 2.286 | 2.742 | 1.969 | 3.818 |
| ΔTrend (year) | 1.219 | 1.079 | 1.379 | 1.032 | 0.882 | 1.207 | 1.454 | 1.200 | 1.762 |
| Past-year quit attempt<sup>4a</sup> |  |  |  |  |  |  |  |  |  |
| Trend (year) | 0.918 | 0.883 | 0.955 | 0.933 | 0.883 | 0.985 | 0.910 | 0.862 | 0.962 |
| Level | 1.417 | 1.297 | 1.547 | 1.349 | 1.193 | 1.527 | 1.460 | 1.290 | 1.654 |
| ΔTrend (year) | 1.074 | 1.016 | 1.136 | 1.018 | 0.940 | 1.102 | 1.115 | 1.031 | 1.206 |
| Number of past-year quit attempts<sup>5a*</sup> |  |  |  |  |  |  |  |  |  |
| Trend (year) | 1.030 | 1.003 | 1.058 | 1.043 | 1.006 | 1.081 | 1.020 | 0.981 | 1.061 |
| Level | 0.970 | 0.911 | 1.032 | 0.893 | 0.821 | 0.971 | 1.034 | 0.944 | 1.133 |
| ΔTrend (year) | 0.997 | 0.958 | 1.038 | 1.023 | 0.969 | 1.081 | 0.981 | 0.926 | 1.040 |
| Medium-term abstinence<sup>6b</sup> |  |  |  |  |  |  |  |  |  |
| Trend (year) | 0.817 | 0.693 | 0.963 | 0.761 | 0.623 | 0.930 | 0.912 | 0.698 | 1.192 |
| Level | 1.575 | 1.101 | 2.254 | 1.600 | 1.008 | 2.539 | 1.479 | 0.852 | 2.566 |
| ΔTrend (year) | 1.424 | 1.169 | 1.735 | 1.571 | 1.228 | 2.009 | 1.240 | 0.905 | 1.699 |

**Table 2. (continued)**
|  | Overall |  |  | Social grades ABC1 |  |  | Social grades C2DE |  |  |
| --- | --- | --- | --- | --- | --- | --- | --- | --- | --- |
|  | RR /<br>*Exp. B*<sup>*</sup> | 95% CI |  | RR /<br>*Exp. B*<sup>*</sup> | 95% CI |  | RR /<br>*Exp. B*<sup>*</sup> | 95% CI |  |
|  |  | Lower | Upper |  | Lower | Upper |  | Lower | Upper |
| Use of prescription medication<sup>5b</sup> |  |  |  |  |  |  |  |  |  |
| Trend (year) | 1.052 | 0.890 | 1.244 | 1.216 | 0.933 | 1.584 | 1.013 | 0.811 | 1.266 |
| Level | 1.286 | 0.887 | 1.865 | 0.769 | 0.415 | 1.403 | 1.499 | 0.925 | 2.428 |
| ΔTrend (year) | 0.845 | 0.662 | 1.080 | 0.860 | 0.580 | 1.275 | 0.826 | 0.601 | 1.135 |
| Use of behavioural support<sup>5b</sup> |  |  |  |  |  |  |  |  |  |
| Trend (year) | 0.838 | 0.696 | 1.009 | 0.790 | 0.608 | 1.026 | 0.876 | 0.670 | 1.143 |
| Level | 2.330 | 1.553 | 3.496 | 2.204 | 1.212 | 4.008 | 2.363 | 1.342 | 4.161 |
| ΔTrend (year) | 1.032 | 0.804 | 1.326 | 1.118 | 0.777 | 1.607 | 0.980 | 0.690 | 1.391 |
| Use of e-cigarettes<sup>5b</sup> |  |  |  |  |  |  |  |  |  |
| Trend (year) | 0.959 | 0.898 | 1.025 | 0.991 | 0.898 | 1.093 | 0.941 | 0.859 | 1.030 |
| Level | 0.788 | 0.666 | 0.932 | 0.711 | 0.552 | 0.915 | 0.841 | 0.670 | 1.056 |
| ΔTrend (year) | 1.232 | 1.111 | 1.365 | 1.241 | 1.061 | 1.452 | 1.225 | 1.067 | 1.408 |

<sup>1</sup> Among all adults. <sup>2</sup> Among 18-24 year-olds. <sup>3</sup> Among 45-65 year-olds. <sup>4</sup> Among past-year smokers. <sup>5</sup> Among past-year smokers who made ≥1 quit attempt in the past 12 months. <sup>6</sup> Among past-year smokers who made ≥1 quit attempt 6-12 months before completing the survey.
<sup>a</sup> Adjusted for seasonality, age, gender, social grade, and region. <sup>b</sup> Adjusted for seasonality, age, gender, social grade, region, cigarette dependence, and national expenditure on tobacco control mass media campaigns.
\* Number of quit attempts was analysed as a continuous variable and was log-transformed for analysis. Results are reported as exponentiated coefficients.
Trend is the underlying secular trend. Level is the step-level change associated with the start of the pandemic.
ΔTrend is the change in trend (slope) following the start of the pandemic.

**Figure 1.**
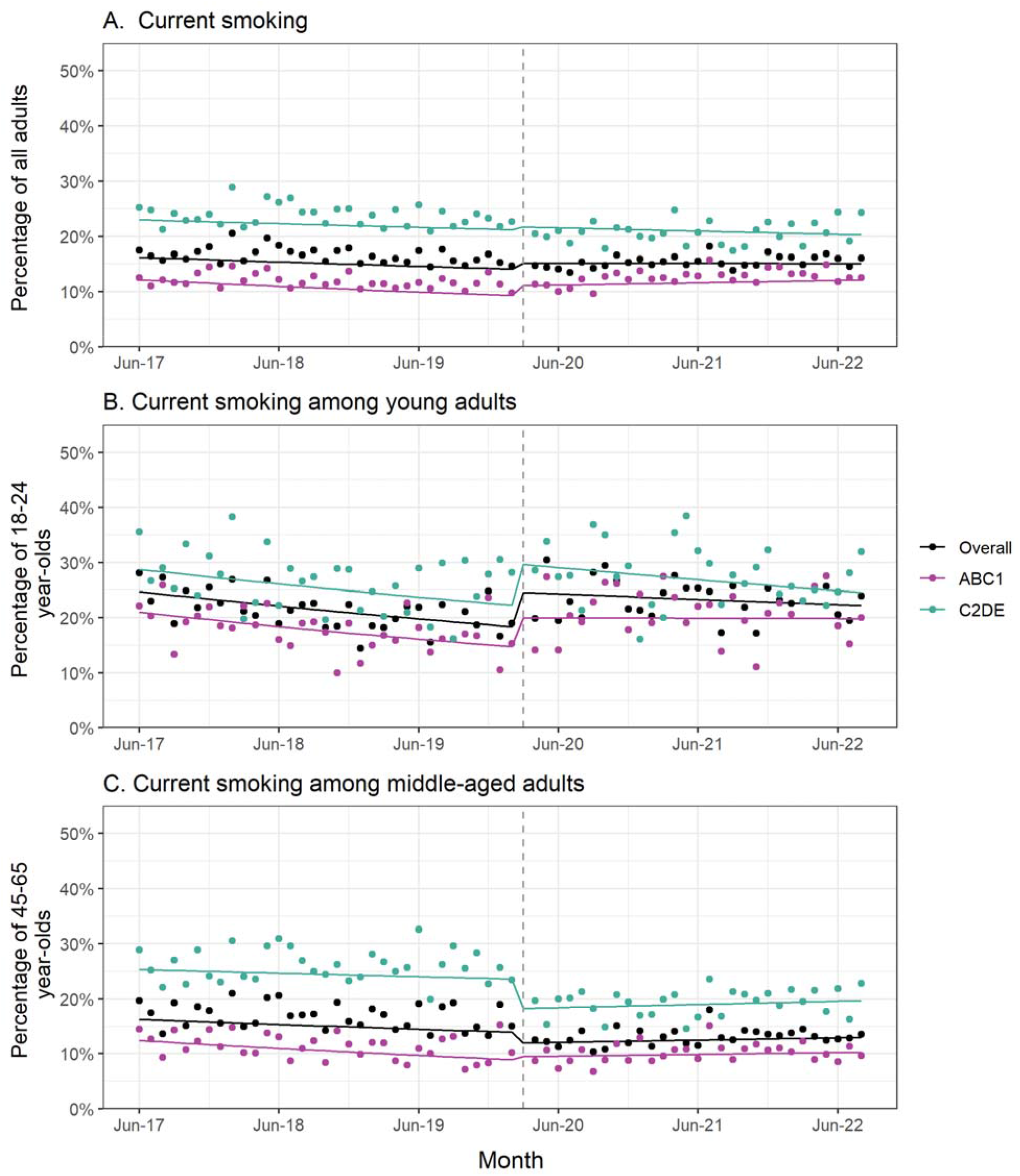
Current smoking, overall and by age and social grade. Panels show trends in the prevalence of current smoking among (A) adults in England (*unweighted n: overall = 101,960, ABC1 = 64,088, C2DE = 37,872*), (B) 18-24 year-olds (*unweighted n: overall = 12,455, ABC1 = 7,766, C2DE = 4,689*), and (C) 45-65 year-olds (*unweighted n: overall = 34,332, ABC1 = 22,401, C2DE = 11,931*), June 2017 to August 2022. Lines represent modelled weighted prevalence over the study period, adjusted for covariates. Points represent raw weighted prevalence by month. The vertical dashed line indicates the timing of the start of the Covid-19 pandemic in England (March 2020).

Overall, among adults in England, the start of the Covid-19 pandemic was associated with a negligible step-level change in current smoking (Figure 1A). However, there was a notable change in trend. Before the pandemic, smoking prevalence fell by 5.2% per year (RR_trend_=0.948); this rate of decline slowed during the pandemic (RR_Δtrend_=1.052, 95%CI=1.014-1.090) to 0.3% per year (RR_trend_ x RR_Δtrend_ = 0.948 × 1.052 = 0.997; Figure 1A). At the start of the pandemic (March 2020), smoking prevalence was estimated at 15.1%. In July 2022, it was virtually unchanged, at 15.0%.

Stratified analyses showed a 20.1% (95%CI=10.1-31.0%) step-level increase in smoking prevalence among adults from social grades ABC1 at the start of the pandemic, followed by a slowing in the pre-pandemic decline to the point where progress in reducing smoking reversed (+3.6% per year compared with -9.5% per year before the pandemic, RR_Δtrend_=1.145, 95%CI=1.083-1.211; Figure 1A). By contrast, there was no increase in smoking prevalence among those from social grades C2DE, and it appeared the modest (∼3% per year) pre-pandemic decline continued (Figure 1A).

When we looked at current smoking in different age groups, we saw divergent changes associated with the pandemic: a 34.9% (95%CI=17.7-54.7%) step-level increase among 18-24 year-olds (Figure 1B) but a 13.6% (95%CI=4.4-21.9%) step-level decrease among 45-65 year-olds (Figure 1C). While the rise in smoking among young adults was similar across social grades, the fall among middle-aged adults was limited to those from social grades C2DE (−22.4%, 95%CI=-10.7 to -32.6%). As we observed overall, progress in reducing smoking stopped among social grades ABC1 during the pandemic (from -12.4% to - 0.3% per year among 18-24 year-olds, RR_Δtrend_=1.138, 95%CI=1.004-1.290; and from -11.7% to +3.4% per year among 45-65 year-olds, RR_Δtrend_=1.171, 95%CI=1.055-1.300) but was similar to pre-pandemic rates within social grades C2DE (Figure 1B and 1C).

The data indicated these changes were sustained over time (Figure 1), rather than short-lived pulse effects during the early months of the pandemic (Supplementary Table 2).

### Quitting activity

Table 2 summarises the GAM results. Figure 2 shows trends in quitting activity over the study period.

**Figure 2.**
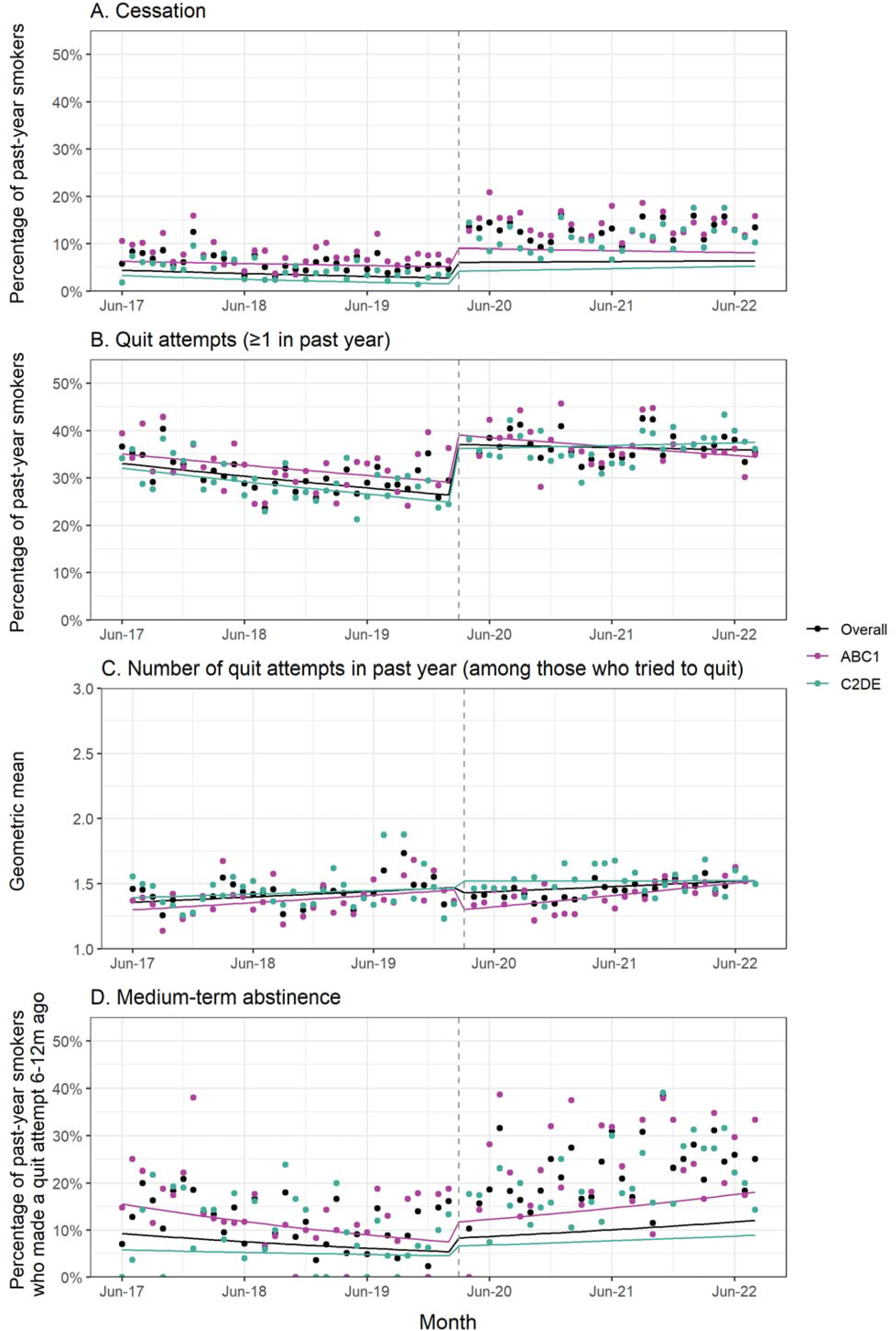
Quitting activity, overall and by social grade. Panels show trends in the prevalence of (A) cessation and (B) making at least one quit attempt in the past year among past-year smokers (*unweighted n: overall = 17,964, ABC1 = 8,802, C2DE = 9,162*), (C) the weighted geometric mean number of past-year quit attempts among past-year smokers who made at least one quit attempt (*unweighted n: overall = 5,754, ABC1 = 2,908, C2DE = 2,846*), and (D) the prevalence of medium-term abstinence among past-year smokers who made a quit attempt 6-12 months ago (*unweighted n: overall = 2,855, ABC1 = 1,489, C2DE = 1,366*), June 2017 to August 2022. Lines represent modelled weighted prevalence (or means) over the study period, adjusted for covariates. Points represent raw weighted prevalence (or means) by month. The vertical dashed line indicates the timing of the start of the Covid-19 pandemic in England (March 2020). Corresponding data without adjustment for dependence are shown in Supplementary Figure 1 and Supplementary Table 3.

Among past-year smokers, the pandemic was associated with a 120.4% (95%CI=79.4-170.9%) step-level increase in cessation (Figure 2A). This increase was similar at 154.4% (95%CI=104.8-216.1%) when cigarette dependence was not adjusted for (Supplementary Table 3, Supplementary Figure 1A) despite mean cigarette dependence only decreasing very slightly during the pandemic (Supplementary Table 4, Supplementary Figure 3). There was also a change in trend: the prevalence of cessation was reducing before the pandemic at a rate of 16.1% per year (RR_trend_=0.839); this rate of decline slowed during the pandemic (RR_Δtrend_=1.219, 95%CI=1.079-1.379) to 2.3% (Figure 2A). The change in trend was driven by social grades C2DE, among whom the rate of cessation was reversed from -24.5% per year before the pandemic to +9.8% per year during the pandemic (RR_Δtrend_=1.454, 95%CI=1.200-1.762; Figure 2A). By contrast, the more modest (7.4%) pre-pandemic decline in cessation among those from social grades ABC1 appeared to continue (Figure 2A).

The pandemic was also associated with a 41.7% (95%CI=29.7-54.7%) step-level increase in the proportion of past-year smokers who made ≥1 quit attempt (Figure 2B). The rate of decline in quit attempts slowed from 8.2% to 1.4% per year (RR_Δtrend_=1.074, 95%CI=1.016-1.136; Figure 2B); again, this was driven by those from social grades C2DE, with no significant change in trend among social grades ABC1 (Figure 2B). Among those who tried to quit, there was little change in the mean number of attempts made (Figure 2C).

Among those who made a quit attempt 6-12 months before the survey, the pandemic was associated with a 57.5% (95%CI=10.1-125.4%) step-level increase in medium-term abstinence (Figure 2D). This increase was 94.9% (95%CI=29.4-193.6%) when cigarette dependence was not adjusted for (Supplementary Table 3, Supplementary Figure 1D). There was also a change in trend: before the pandemic, the prevalence of medium-term abstinence declined by 18.3% per year but this trend reversed (RR_Δtrend_=1.424, 95%CI=1.169-1.735) and increased by 16.3% per year during the pandemic (Figure 2D). The step-level increase and change in trend were only statistically significant among those from social grades ABC1, but the pattern of results was similar for social grades C2DE (Figure 1D). The relatively small sample sizes for this outcome meant that estimates were imprecise.

While analyses of pulse effects showed increases in quitting activity in the first 2-3 months of the pandemic (Supplementary Table 2), it is clear from Figure 2 and the change in trend results (Table 2) that these increases were sustained through to August 2022.

### Use of cessation support

Table 2 summarises the GAM results. Figure 3 shows trends in use of cessation support over the study period.

**Figure 3.**
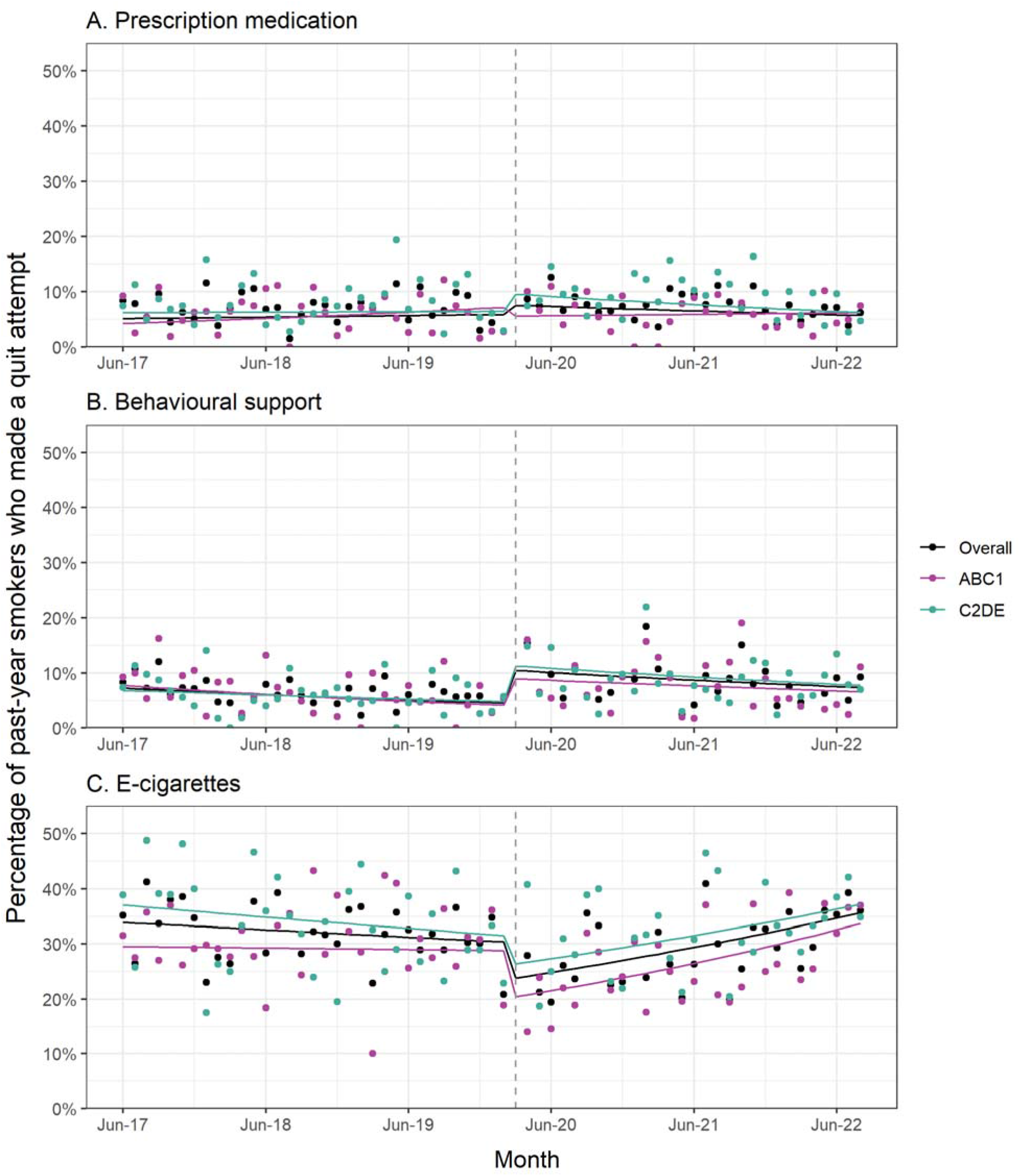
Use of support by smokers in quit attempts, overall and by social grade. Panels show trends in the prevalence of use of (A) prescription medication, (B) behavioural support, and (C) e-cigarettes in the most recent quit attempt among past-year smokers who made a least one quit attempt (*unweighted n: overall = 5,754, ABC1 = 2,908, C2DE = 2,846*), June 2017 to August 2022. Lines represent modelled weighted prevalence over the study period, adjusted for covariates. Points represent raw weighted prevalence by month. The vertical dashed line indicates the timing of the start of the Covid-19 pandemic in England (March 2020). Corresponding data without adjustment for dependence are shown in Supplementary Figure 2 and Supplementary Table 3.

Among past-year smokers who made a quit attempt, the start of the Covid-19 pandemic was associated with little change in the use of prescription medication (Figure 3A). Point estimates for a step-level change were in opposite directions for those from social grades ABC1 and C2DE, but neither group had a statistically significant change. This finding was robust to the exclusion of varenicline from this variable (Supplementary Table 5).

However, the pandemic was associated with changes in the use of behavioural support and e-cigarettes for quitting smoking. There was a 133.0% (95%CI=55.3-249.6%) step-level increase in use of behavioural support, followed by a continuation of the modest pre-pandemic decline (Figure 3B). By contrast, there was a 21.2% (95%CI=6.8-33.4%) step-level decrease in use of e-cigarettes (Figure 3C). This change was short-lived (Supplementary Table 2) because there was also a change in trend, reversing this step-level decline: before the pandemic, the proportion of smokers using e-cigarettes in a quit attempt fell by 4.1% per year; during the pandemic, it increased by 18.1% per year (RR_Δtrend_=1.232, 95%CI=1.111-1.365, Figure 3C). These changes were similar across social grades.

Changes in use of cessation support were similar when cigarette dependence was not adjusted for (Supplementary Table 3, Supplementary Figure 2).

## Discussion

Before the Covid-19 pandemic, smoking prevalence had been falling among adults in England at a near linear rate for more than 20 years.^14^ Our data show that this decline almost completed stopped since the pandemic began, resulting from changes in smoking and quitting behaviours. There were sustained changes in smoking prevalence in different age groups: an increase among 18-24 year-olds, indicating a potential rise in the uptake of smoking, offset by a decrease among 45-65 year-olds, which also suggested no evidence of a substantial rise in late relapse. Among more advantaged social grades, the pre-pandemic decline halted; among the less advantaged, there was no change in the pre-pandemic trend, suggesting that smoking inequalities have narrowed. There were sustained increases in quitting activity among past-year smokers: cessation rates more than doubled during the pandemic, and there were also increases in rates of quit attempts and medium-term abstinence. Among smokers from more disadvantaged social grades, declining pre-pandemic trends in cessation and quit attempts flattened or reversed. Among those who tried to quit, there was little change in the number of attempts made or in the use of prescription medication in a quit attempt, but there was a short-term rise in the use of behavioural support and a short-term fall in the use of e-cigarettes, which recovered during the pandemic period.

These results build upon and extend our previous analysis of changes in smoking and quitting during the first Covid-19 lockdown in England (April–July 2020).^1^ We applied more complex modelling techniques to data collected over a longer period (up to August 2022), offering insight into the longer-term impact of the pandemic on smoking. In addition, we analysed additional outcomes (e.g., relapse) to provide a more complete picture of the pandemic’s impacts. There are three key findings.

First, the pandemic has been associated with opposing changes in smoking behaviour: the proportion of smokers who have tried to quit and succeeded has risen, but so has the proportion of young adults who smoke. In the absence of any adverse effect on relapse, the net result is stable overall smoking prevalence, which appears to have halted years of steady decline in prevalence pre-pandemic. As we have discussed previously,^10^ the unique circumstances brought about by the pandemic may have prompted some smokers to quit (e.g., by providing a ‘teachable moment’, disrupting daily routines, or reducing social smoking cues) while encouraging other people to start smoking (e.g., to relieve stress or boredom). In particular, younger adults have experienced higher levels of stress, upheaval, and social isolation during the pandemic,^15,16^ which might have contributed to increased smoking prevalence in this group. Based on the stagnation in the decline of smoking prevalence in the two years since the pandemic began, there is an urgent need for bold policy action. Even based on pre-pandemic trends, the UK Government’s smokefree 2030 target would not have been achieved until at least 2037.^17^ The stagnation appears to have been caused by an increase in smoking among young adults. There is a need to understand the motives driving young adults to take up smoking to develop interventions and public health messaging to combat this. Increasing the age of sale of cigarettes is one strategy that may be effective both in reducing uptake and in narrowing inequalities in smoking.^18,19^

Second, the pandemic appears to have had equity-positive impacts on smoking. Progress in reducing smoking prevalence has historically been slower for disadvantaged groups.^20,21^ However, while the (considerable) pre-pandemic decline in smoking prevalence among social grades ABC1 levelled off during the pandemic, the more modest decline among social grades C2DE continued. While it is encouraging to see inequalities in smoking narrow during the pandemic, ideally this would be the result of accelerating reductions in smoking prevalence among disadvantaged groups, rather than slowing the decline among advantaged groups. Trends in quitting activity also showed evidence of narrowing inequalities: a consistent decline among social grades ABC1 but a levelling off during the pandemic among social grades C2DE. Possible explanations for these differences include those from social grades C2DE being more likely to (i) experience financial impacts of the pandemic (e.g., due to job loss or reduced earnings) which make (taking up or continuing) smoking less affordable, or (ii) work in front-line jobs that increase exposure to Covid-19 and might make quitting smoking higher priority.^22–24^ In working toward the smokefree 2030 target, there is a need for action to reignite progress in reducing smoking among the more advantaged social grades and identify ways to accelerate the decline among less advantaged groups.

Finally, the pandemic has not had an enduring impact on the use of evidence-based support by smokers trying to quit. At the start of the pandemic, there was a fall in the use of e-cigarettes, the most popular quitting aid used by smokers in England.^13^ It is possible this resulted from unsubstantiated concerns that vaping might exacerbate the risk of contracting or experiencing complications from Covid-19,^25^ or difficulties in getting to vape shops before businesses pivoted, which were not exempted from lockdown rules. The decline in e-cigarette use was offset by a rise in the use of behavioural support (e.g., stop smoking services or digital support via websites or apps), so there was no adverse impact on the success of quit attempts (as indicated by increased rates of cessation and medium-term abstinence). Over the longer term, use of different types of support returned to pre-pandemic levels. Given most smokers who tried to quit during the pandemic did not report using any evidence-based support, there remains substantial opportunity to boost success in quitting by directing smokers to effective support. Increased investment in national tobacco control mass media campaigns may be a cost-effective means to achieve this.^26,27^

Strengths of this study include the large, representative sample, the repeat cross-sectional design with data pre-dating the pandemic, and the broad range of data captured on smoking and quitting behaviour. There were also limitations. There is no direct assessment of late relapse in the Smoking Toolkit Study. We therefore analysed changes in smoking prevalence among 45-65 year-olds as a proxy variable on the basis that any increase in this age group would most likely be driven by a rise in late relapse rather than uptake. However, it is possible that any small increase in late relapse among this group was offset by more people quitting.

Another limitation was the change in modality of data collection from face-to-face (before the pandemic) to telephone interviews (during the pandemic). While this was unavoidable due to social distancing restrictions, it is possible that it contributed to some of the changes we observed — especially step-level changes, which would be most sensitive to any effects of the switch. Nevertheless, comparisons of the face-to-face and telephone data within the Smoking Toolkit Study,^11^ combined with previous studies showing a high degree of comparability between face-to-face and telephone interviews,^28,29^ suggest that it is reasonable to compare data collected via the two methods. Given there were no further updates to methods after April 2020, changes in the slope of trends are unlikely to be explained by the switch in methodology.

In conclusion, the rate of decline in adult smoking prevalence in England has stagnated during the Covid-19 pandemic. Reductions in smoking prevalence among middle-aged adults and sustained increases in quit attempts and cessation among smokers have been offset by a sustained rise in uptake among young adults. Changes in use of support predominantly occurred in the early stages of the pandemic and have since returned to usual levels. There was no evidence to suggest the pandemic increased the risk of early or late relapse. The slowing in the rate of decline in smoking prevalence was pronounced in more advantaged social grades.

## Data Availability

Data and code are available from the corresponding author on reasonable request.

## Declarations

### Ethics approval and consent to participate

Ethical approval for the STS was granted originally by the UCL Ethics Committee (ID 0498/001). The data are not collected by UCL and are anonymised when received by UCL.

### Data sharing

Data and code are available from the corresponding author on reasonable request.

### Author contributions

Conceptualisation: SJ, HTB, LS, EB, JB. Data curation: JB. Formal analysis: SJ, HTB. Funding acquisition: LS, JB. Investigation: SJ, HTB, LS, EB, JB. Methodology: SJ, HTB, LS, EB, JB. Supervision: JB. Visualisation: SJ, HTB. Writing – original draft: SJ. Writing – review & editing: HTB, LS, EB, JB.

