## Supplementary material for "Have there been sustained impacts of the Covid-19 pandemic on trends in smoking prevalence, uptake, quitting, use of treatment, and relapse? A monthly population study in England, 2017-2022"

**Part 1: Detailed literature review**

**Part 2: Full details of measures used**

**Part 3: Unweighted descriptive statistics**

| **Supplementary Table 1**. | Unweighted descriptive statistics |
| --- | --- |

**Part 4: Sensitivity analyses**

| *1. Pulse effects* | |
| --- | --- |
| **Supplementary Table 2**. | Sensitivity analysis: pulse effects |
| *2. Excluding dependence as a covariate* | |
| **Supplementary Table 3**. | Sensitivity analysis: excluding cigarette dependence as a covariate |
| **Supplementary Figure 1.** | Quitting activity, with and without adjustment for dependence |
| **Supplementary Figure 2.** | Use of support by smokers in quit attempts, with and without adjustment for dependence |
| *3. Changes in dependence* | |
| **Supplementary Table 4.** | Associations between the Covid-19 pandemic and cigarette dependence |
| **Supplementary Figure 3.** | Cigarette dependence |
| *4. Excluding varenicline from prescription medication* | |
| **Supplementary Table 5.** | Sensitivity analysis: excluding varenicline from analysis of use of prescription medication |

### Part 1: Detailed literature review

The extent to which the Covid-19 pandemic has affected ***smoking prevalence*** remains unclear. A meta-analysis of international studies up to November 2020 reporting on changes in smoking behaviour after the onset of the pandemic indicated there had been a significant decline in smoking prevalence (12 studies; prevalence ratio=0.87, 95%CI [confidence interval]=0.79-0.97).^2^ However, there was a very high degree of heterogeneity across studies (I^2^=99.3%), with some reporting a significant increase in prevalence, some a significant decrease, and others no change. In addition, just two studies were representative, with the majority using convenience samples recruited via social media or other online platforms, and almost all were judged to be at high risk of bias. Further data on changes in smoking prevalence were published in 2021. In the UK, the Annual Population Survey (a large, representative household survey) showed a large but implausible decline in the proportion of adults who smoked cigarettes, from 13.8% in the first quarter of 2020 to 12.3% in April to December of 2020. The magnitude of the decline may have been partially attributable to the change in mode of data collection from predominantly face-to-face interviews to telephone-only.^4^ The Opinions and Lifestyle Survey, another representative survey which was already remote at the start of the pandemic (telephone/online), suggested smoking prevalence increased at the start of the pandemic, from 15.3% in April 2020 to a peak of 16.3% in August 2020 (although this change was not statistically significant), before falling significantly to 12.5% in December 2020.^4^ The Smoking Toolkit Study (a representative monthly survey) showed no overall change in smoking prevalence among adults in England during the first Covid-19 lockdown in England (despite a change from face-to-face to telephone interviews), but detected a large increase in smoking prevalence among young adults (18-34 years; from 21.5% between August 2019 and February 2020 to 26.8% between April and July 2020).^1^

Data on changes in ***uptake*** of smoking during the Covid-19 pandemic are scarce. Sarich et al.’s meta-analysis^2^ identified four studies that provided data on the proportion of smokers who started or restarted smoking during the pandemic; the pooled estimate was 2%, with a high degree of heterogeneity between studies (I^2^=91.7%). All the studies used convenience sampling and were judged at high risk of bias. A survey of young people in Great Britain by Action on Smoking and Health showed no evidence of an increase in uptake (indexed by ever-use of cigarettes) among 11-17 year-olds.^5^ Analysing changes in smoking prevalence among young adults could offer insight into any impact of the pandemic on uptake, as increases in prevalence in this age group would likely be largely driven by uptake rather than relapse.

More data are available on ***quitting***, with studies indicating the pandemic may have prompted many smokers to stop. The Smoking Toolkit Study showed substantial increases in the overall rate of cessation among smokers (from 3.9% in August 2019 to February 2020 to 10.0% in April to July 2020) and the success rate of quit attempts (from 12.7% to 25.3%).^1^ The proportion of younger smokers (18-34 years) who reported making a serious attempt to quit increased by 39.9% (from 32.1% to 44.9%).^1^ The ITC Four Country survey conducted in April-June 2020 found almost half (46.7%) of current smokers had thought about quitting because of Covid-19.^3^ Roughly equal proportions reported having reduced (14.2%) or increased (14.6%) the amount they smoked since the coronavirus outbreak. Smokers who reported positive behaviour change (quit attempt/reduction) tended to be less dependent, more concerned about personal susceptibility to infection, and were more likely to believe that Covid-19 is more severe for smokers. There was substantial between-country variation: smokers in Australia were less likely than those in England, Canada, or the United States to have tried to quit or reduced their smoking, which may be attributable to the significantly lower impact of Covid-19 on Australia during the early stages of the pandemic. It is not clear whether the pandemic has had an influence on the frequency of quit attempts.

There is limited evidence on changes in ***use of cessation support*** since the start of the pandemic among smokers who have tried to quit. The Smoking Toolkit Study showed an increase in the proportion of smokers making a quit attempt using remote support (telephone support/websites/apps; from 2.4% in August 2019 to February 2020 to 6.8% in April to July 2020) and a decline in the proportion using evidence-based support (defined as face-to-face behavioural support, prescription medication [varenicline, bupropion, or nicotine replacement therapy (NRT)], e-cigarettes, or over-the-counter NRT; from 53.8% to 44.8%), but these changes were not statistically significant over and above changes over the same period a year earlier.^1^ These analyses were limited by the small number of smokers reporting a past-year quit attempt,^1^ so cannot be considered conclusive.

Finally, relatively little is known about what impact the pandemic has had on ***relapse*** to smoking among ex-smokers. Given studies have generally observed large increases in quitting activity^1,3^ but little or no reduction in smoking prevalence,^1,2,4^ it is possible that increased rates of cessation are being offset by increased rates of relapse. Examining changes in smoking prevalence among middle-aged and older adults could offer insight into the impact of the pandemic on late relapse, as any increases in smoking prevalence in this age group would likely be largely driven by relapse rather than uptake (as few people take up smoking after the age of 25^6^).

### Part 2: Full details of all measures used

#### Outcome variables

We assessed the following smoking outcomes:

- Among all adults: current smoking;
- Among 18-24 year-olds: uptake of smoking (indexed by current smoking);
- Among 45-65 year-olds: late relapse (indexed by current smoking);
- Among past-year smokers: cessation, quit attempts;
- Among past-year smokers who made ≥1 quit attempt in the past 12 months: number of quit attempts made, use of prescription medication, use of behavioural support, use of e-cigarettes;
- Among past-year smokers who made ≥1 quit attempt 6-12 months before completing the survey: medium-term abstinence.

Smoking status was assessed with the question: ‘Which of the following best applies to you? (a) I smoke cigarettes (including hand-rolled) every day; (b) I smoke cigarettes (including hand-rolled), but not every day; (c) I do not smoke cigarettes at all, but I do smoke tobacco of some kind (e.g. pipe, cigar or shisha); (d) I have stopped smoking completely in the last year; (e) I stopped smoking completely more than a year ago; (f) I have never been a smoker (i.e. smoked for a year or more)’. ***Current smoking*** was coded 1 for those who reported smoking any type of tobacco [i.e. responses (a–c)] and 0 for those who reported being a former or never smoker [responses (d–f)]. Past-year smoking was coded 1 for those who reported current smoking or having stopped in the past year [responses (a–d)] and 0 for those who reported stopping more than a year ago or never smoking [responses (e–f)].

***Uptake*** was assessed by current smoking prevalence among 18-24 year olds (because any increases in this age group would largely be driven by uptake rather than relapse).

***Late relapse*** was assessed by current smoking prevalence among 45-65 year olds (because an increase in this age group would largely be driven by relapse rather than uptake). The upper age limit for this group was selected to minimise any impact of increased mortality at older ages during the pandemic on smoking prevalence.

Among past-year smokers, ***cessation*** was coded 1 for those who reported having stopped smoking completely in the last year [response (d) to the measure of smoking status described above] and 0 for those who reported being a current smoker [responses (a–c)].

Among past-year smokers, quit attempts were assessed with the question: ‘How many serious attempts to stop smoking have you made in the last 12 months? By serious attempt I mean you decided that you would try to make sure you never smoked again. Please include any attempt that you are currently making and please include any successful attempt made within the last year’. Those who reported ***making ≥1 serious quit attempt*** in the past year were coded 1; otherwise they were coded 0.

Among those who reported making ≥1 quit attempt, ***number of past-year quit attempts*** was analysed as a continuous variable. We log-transformed the number of past-year quit attempts variable for analysis to normalise the skewed distribution and reported results as geometric means for ease of interpretation.

Among those who reported making ≥1 quit attempt, use of cessation support in the most recent quit attempt was also assessed with the question: ‘Which, if any, of the following did you try to help you stop smoking during the most recent serious quit attempt?’. Participants were asked to indicate all that apply.

- ***Use of prescription medication*** was coded 1 for those who reported using varenicline, bupropion, or NRT on prescription, and 0 for those who did not report using any of these. Because varenicline was recalled in 2021 by its manufacturer, Pfizer Ltd, we ran a sensitivity analysis excluding varenicline to check that any apparent impact of the pandemic on use of prescription medication was not driven by an artefactual decline in use of varenicline due to unavailability.
- ***Use of behavioural support*** was coded 1 for those who reported attending a stop smoking group or one or more stop smoking one-to-one counselling/advice/support session(s) (either in person or remotely) or using telephone support, a website, an app, or written self-help materials, and 0 for those who did not report using any of these.
- ***Use of e-cigarettes*** was coded 1 for those who reported using e-cigarettes, and 0 for those who did not.

Among past-year smokers who reported making ≥1 quit attempt in the 6-12 months before completing the survey, ***medium-term abstinence*** following the most recent quit attempt in this period was coded 1 for those who did not report a subsequent quit attempt and responded that they were ‘still not smoking’ to the question ‘How long did your most recent serious quit attempt last before you went back to smoking?’. Medium-term abstinence was coded 0 for participants who reported a making a subsequent quit attempt (i.e., any quit attempt in the 0-6 months prior to the survey) or reported being a current smoker.

#### Trends, pandemic effects, and seasonality

Time was measured in months throughout the study period (coded 1…*n*, where *n* was the total number of waves), which allowed us to account for ***underlying secular trends***. An additional variable reflected the pre-pandemic and pandemic periods in order to identify ***step-level changes*** in the outcomes (coded 0 before the start of the pandemic [June 2017 through February 2020] and 1 after [April 2020 through August 2022]). Finally, we included a variable to reflect the ***change in trend*** (slope) following the start of the pandemic (coded 0 before the pandemic and 1…*m* from April 2020 onwards, where *m* was the number of waves after the start of the pandemic). We controlled for ***seasonality*** (month-of-year effects) using cyclic cubic splines (see Statistical Analysis).

#### Covariates

Analyses of smoking prevalence, uptake, late relapse, and quit attempts were adjusted for age, gender, social grade and region. ***Age*** was analysed as a continuous variable. ***Gender*** was self-reported as ‘man’, ‘woman’, or ‘in another way’. Due to low numbers, those who described their gender ‘in another way’ were excluded from analyses that adjusted for gender. ***Social grade*** was categorised as ABC1 (which includes managerial, professional and intermediate occupations) versus C2DE (which includes small employers and own-account workers, lower supervisory and technical occupations, and semi-routine and routine occupations, never workers and long-term unemployed). ***Region*** in England was categorised as London, South, Central, and North.

Analyses of smoking cessation, medium-term abstinence, and use of support were adjusted for these sociodemographic characteristics, level of dependence (because more dependent smokers tend to be less likely to quit and more likely to use support^1^), and government spending on tobacco control mass media campaigns.^2–4^ ***Cigarette dependence*** was assessed with self-reported ratings of strength of urges to smoke over the past 24 hours [not at all (coded 0), slight (1), moderate (2), strong (3), very strong (4) and extremely strong (5)]. This variable was also coded ‘0’ for smokers who responded ‘not at all’ to the (separate) question: ‘How much of the time have you spent with the urge to smoke?’^5^ Monthly ***expenditure on tobacco control mass media campaigns*** (in £) was obtained from the Office for Health Improvement and Disparities and adjusted for inflation. While there were other potentially relevant population-level covariates, such as affordability of e-cigarettes or prevalence of e-cigarette use, we did not adjust for them in this study as they could plausibly have been affected by the pandemic.^6^

### Part 3: Unweighted descriptive statistics

**Supplementary Table 1**. Unweighted descriptive statistics

|  | Overall | Before pandemic  (June 2017-Feb 2020) | During pandemic  (April 2020-Aug 2022) |
| --- | --- | --- | --- |
| *N* | *101 960* | *55 349* | *46 611* |
| Age (years) | 50.2 (19.3) | 49.8 (19.7) | 50.7 (18.7) |
| 18-24 | 12.2% | 13.5% | 10.6% |
| 25-44 | 28.5% | 28.5% | 28.5% |
| 45-65 | 33.7% | 32.0% | 35.7% |
| >65 | 25.6% | 25.9% | 25.2% |
| Gender |  |  |  |
| Men | 48.9% | 49.9% | 47.8% |
| Women | 50.8% | 50.0% | 51.6% |
| In another way | 0.3% | 0.1% | 0.6% |
| Social grade C2DE | 37.1% | 39.1% | 34.8% |
| Region in England |  |  |  |
| London | 15.8% | 16.0% | 15.6% |
| South | 25.1% | 23.7% | 26.6% |
| Central | 30.5% | 30.9% | 30.0% |
| North | 28.6% | 29.3% | 27.8% |
| Current smoker | 16.0% | 16.5% | 15.4% |
| Past-year smoker | 17.6% | 17.6% | 17.6% |
| Uptake of smoking^1c^ | 22.0% | 21.2% | 23.3% |
| Late relapse^2c^ | 15.0% | 16.9% | 13.1% |
| Cigarette dependence^3a^ | 1.70 (1.19) | 1.76 (1.14) | 1.62 (1.26) |
| Quit in past year^3^ | 9.2% | 6.1% | 12.8% |
| Tried to quit in past year^3^ | 33.4% | 30.6% | 36.8% |
| Number of past-year quit attempts^4b^ | 1.43 (1.79) | 1.40 (1.77) | 1.45 (1.79) |
| Medium-term abstinence^5^ | 17.2% | 11.9% | 22.2% |
| Use of cessation support^4^ |  |  |  |
| Prescription medication | 7.2% | 7.0% | 7.5% |
| Behavioural support | 7.4% | 6.4% | 8.3% |
| E-cigarettes | 30.6% | 32.2% | 29.1% |
| Monthly inflation-adjusted national tobacco control expenditure (£) | 137 000  (247 000) | 142 000  (237 000) | 131 000  (261 000) |
| Data are shown as percentages or mean (SD), unless otherwise specified.  ^1^ Among 18-24 year olds (*n = 12,455*). ^2^ Among 45-65 year olds (*n = 34,332*). ^3^ Among past-year smokers (*n = 17,964*). ^4^ Among past-year smokers who tried to quit in the past year (*n = 5,754*). ^5^ Among past-year smokers who made a quit attempt that started 6-12 months before completing the survey (*n = 2,855*).  ^a^ Strength of urges to smoke rated on a scale from 0 (none) to 5 (extremely strong).  ^b^ Geometric mean.  ^c^ Indexed by current smoking.  There was a small amount of missing data for some variables (<0.1% gender, 0.1% medium-term abstinence, 2.4% cigarette dependence, 4.1% tried to quit in past year); valid percentages are shown. | | | |

### Part 4: Sensitivity analyses

**Supplementary Table 2**. Sensitivity analysis: pulse effects

|  | | **RR / *Exp.* *B*^*^** | **95% CI** | |
| --- | --- | --- | --- | --- |
|  | |  | **Lower** | **Upper** |
| Current smoking^1a^ | |  |  |  |
|  | 2-month pulse | 1.029 | 0.956 | 1.108 |
|  | 3-month pulse | 0.998 | 0.936 | 1.063 |
| Uptake of smoking^4a^ | |  |  |  |
|  | 2-month pulse | 1.152 | 0.969 | 1.370 |
|  | 3-month pulse | 1.059 | 0.907 | 1.236 |
| Late relapse^6a^ | |  |  |  |
|  | 2-month pulse | 0.932 | 0.807 | 1.076 |
|  | 3-month pulse | 0.894 | 0.792 | 1.009 |
| Cessation^2b^ | |  |  |  |
|  | 2-month pulse | 1.392 | 1.129 | 1.717 |
|  | 3-month pulse | 1.382 | 1.162 | 1.644 |
| Past-year quit attempt^2a^ | |  |  |  |
|  | 2-month pulse | 1.113 | 1.001 | 1.238 |
|  | 3-month pulse | 1.130 | 1.034 | 1.235 |
| Number of past-year quit attempts^3a*^ | |  |  |  |
|  | 2-month pulse | 0.997 | 0.922 | 1.078 |
|  | 3-month pulse | 1.000 | 0.937 | 1.067 |
| Medium-term abstinence^5b^ | |  |  |  |
|  | 2-month pulse | 0.762 | 0.498 | 1.167 |
|  | 3-month pulse | 0.701 | 0.498 | 0.987 |
| Use of prescription medication^3b^ | |  |  |  |
|  | 2-month pulse | 1.019 | 0.614 | 1.693 |
|  | 3-month pulse | 1.349 | 0.943 | 1.930 |
| Use of behavioural support^3b^ | |  |  |  |
|  | 2-month pulse | 2.081 | 1.317 | 3.288 |
|  | 3-month pulse | 1.866 | 1.296 | 2.685 |
| Use of e-cigarettes^3b^ | |  |  |  |
|  | 2-month pulse | 0.743 | 0.584 | 0.944 |
|  | 3-month pulse | 0.706 | 0.575 | 0.867 |

^1^ Among all adults. ^2^ Among past-year smokers. ^3^ Among past-year smokers who made ≥1 quit attempt in the past 12 months. ^4^ Among 18-24 year-olds (indexed by current smoking prevalence). ^5^ Among past-year smokers who made ≥1 quit attempt 6-12 months before completing the survey. ^6^ Among 45-65 year-olds (indexed by current smoking prevalence).

^a^ Adjusted for underlying trend, seasonality, age, gender, social grade, and region. ^b^ Adjusted for underlying trend, seasonality, age, gender, social grade, region, cigarette dependence, and national expenditure on tobacco control mass media campaigns.

* Number of quit attempts was analysed as a continuous variable and was log-transformed for analysis. Results are reported as exponentiated coefficients.

**Supplementary Table 3**. Sensitivity analysis: excluding cigarette dependence as a covariate

|  | | **RR** | **95% CI** | |
| --- | --- | --- | --- | --- |
|  | |  | **Lower** | **Upper** |
| Cessation^1a^ | |  |  |  |
|  | Trend (year) | 0.828 | 0.748 | 0.918 |
|  | Level | 2.544 | 2.048 | 3.161 |
|  | ΔTrend (year) | 1.246 | 1.093 | 1.421 |
| Medium-term abstinence^3b^ | |  |  |  |
|  | Trend (year) | 0.813 | 0.673 | 0.982 |
|  | Level | 1.949 | 1.294 | 2.936 |
|  | ΔTrend (year) | 1.501 | 1.188 | 1.896 |
| Use of prescription medication^2b^ | |  |  |  |
|  | Trend (year) | 1.058 | 0.892 | 1.253 |
|  | Level | 1.189 | 0.816 | 1.735 |
|  | ΔTrend (year) | 0.850 | 0.666 | 1.086 |
| Use of behavioural support^2b^ | |  |  |  |
|  | Trend (year) | 0.836 | 0.694 | 1.005 |
|  | Level | 2.270 | 1.515 | 3.401 |
|  | ΔTrend (year) | 1.036 | 0.808 | 1.330 |
| Use of e-cigarettes^2b^ | |  |  |  |
|  | Trend (year) | 0.960 | 0.898 | 1.026 |
|  | Level | 0.776 | 0.657 | 0.917 |
|  | ΔTrend (year) | 1.232 | 1.112 | 1.365 |

^1^ Among past-year smokers. ^2^ Among past-year smokers who made ≥1 quit attempt in the past 12 months. ^3^ Among past-year smokers who made ≥1 quit attempt 6-12 months before completing the survey.

^a^ Adjusted for seasonality, age, gender, social grade, region, and national expenditure on tobacco control mass media campaigns.

Trend is the underlying secular trend. Level is the step-level change associated with the start of the pandemic. ΔTrend is the change in trend (slope) following the start of the pandemic.

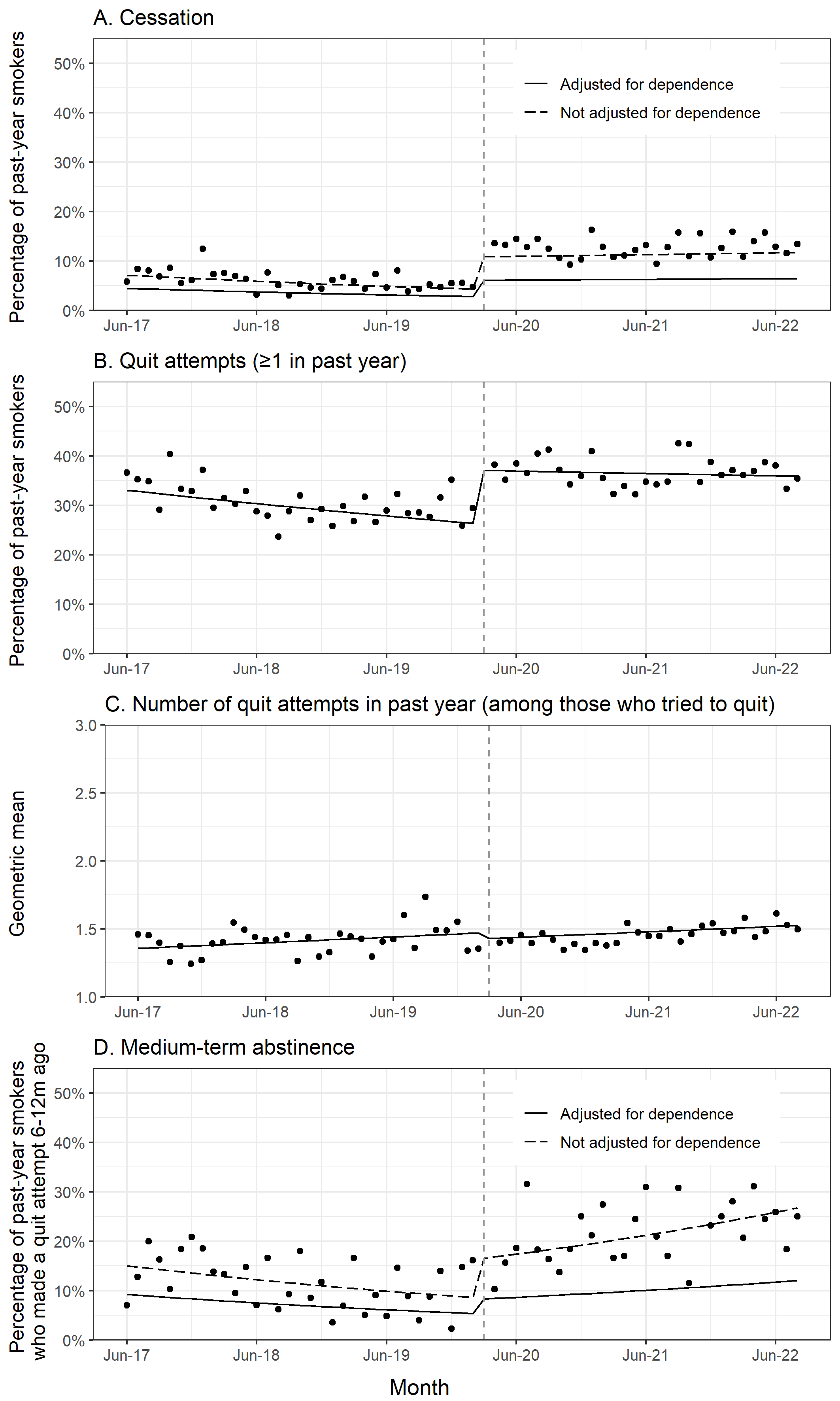

**Supplementary Figure 1. Quitting activity, with and without adjustment for dependence.** Panels show trends in the prevalence of (A) cessation and (B) making at least one quit attempt in the past year among past-year smokers (*unweighted n = 17,964*), (C) the weighted geometric mean number of past-year quit attempts among past-year smokers who made at least one quit attempt (*unweighted n = 5,754*), and (D) the prevalence of medium-term abstinence among past-year smokers who made a quit attempt 6-12 months ago (*unweighted n = 2,855*), June 2017 to August 2022. Lines represent modelled weighted prevalence (or means) over the study period, adjusted for covariates. Points represent raw weighted prevalence (or means) by month. The vertical dashed line indicates the timing of the start of the Covid-19 pandemic in England (March 2020).

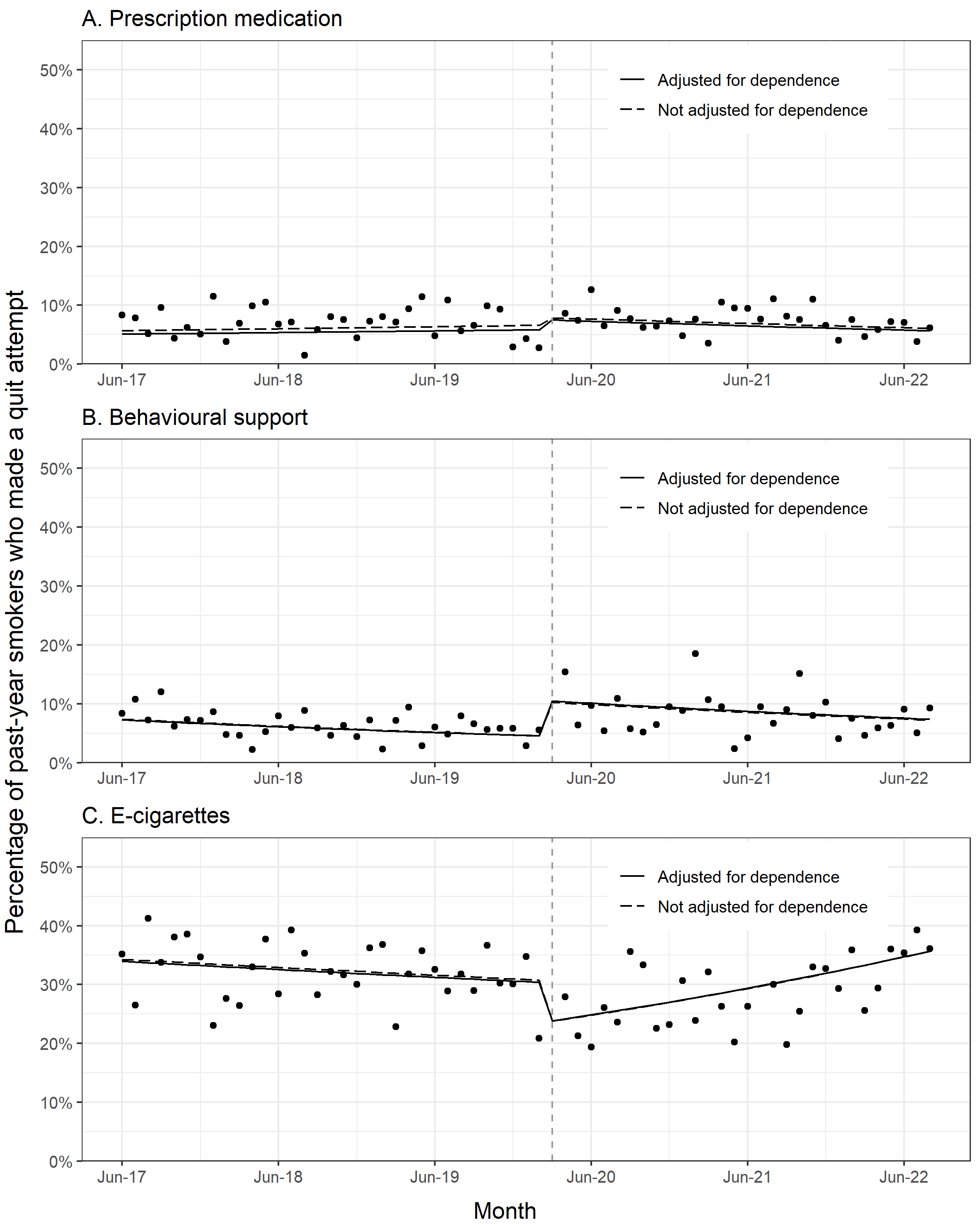

**Supplementary Figure 2. Use of support by smokers in quit attempts, with and without adjustment for dependence.** Panels show trends in the prevalence of use of (A) prescription medication, (B) behavioural support, and (C) e-cigarettes in the most recent quit attempt among past-year smokers who made a least one quit attempt (*unweighted n = 5,754*), June 2017 to August 2022. Lines represent modelled weighted prevalence over the study period, adjusted for covariates. Points represent raw weighted prevalence by month. The vertical dashed line indicates the timing of the start of the Covid-19 pandemic in England (March 2020).

**Supplementary Table 4**. Associations between the Covid-19 pandemic and cigarette dependence

|  | | ***B*** | **95% CI** | |
| --- | --- | --- | --- | --- |
|  | |  | **Lower** | **Upper** |
| Cigarette dependence^1a^ | |  |  |  |
|  | Trend (year) | -0.003 | -0.033 | 0.027 |
|  | Level | -0.107 | -0.179 | -0.035 |
|  | ΔTrend (year) | 0.002 | -0.046 | 0.049 |

^1^ Among past-year smokers.

^a^ Adjusted for seasonality, age, gender, social grade, and region.

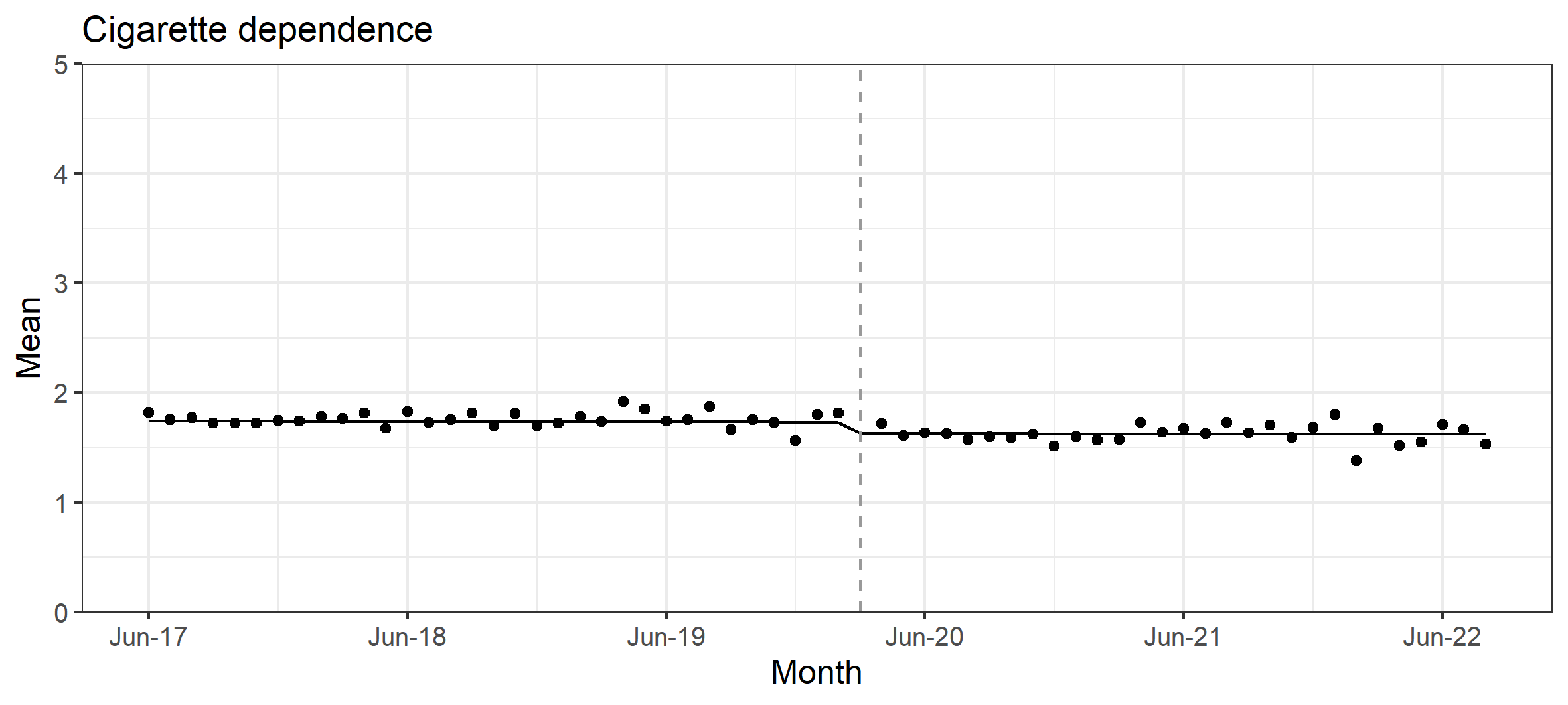

Supplementary Figure 3. Cigarette dependence. Panels show trends in mean cigarette dependence among past-year smokers (*unweighted n = 17,964*), June 2017 to August 2022. Lines represent the modelled weighted mean over the study period, adjusted for covariates. Points represent raw weighted means by month. The vertical dashed line indicates the timing of the start of the Covid-19 pandemic in England (March 2020).

**Supplementary Table 5**. Sensitivity analysis: excluding varenicline from analysis of use of prescription medication

|  | | **RR** | **95% CI** | |
| --- | --- | --- | --- | --- |
|  | |  | **Lower** | **Upper** |
| Use of prescription medication^1a^ | |  |  |  |
|  | Trend (year) | 1.086 | 0.856 | 1.379 |
|  | Level | 0.962 | 0.565 | 1.637 |
|  | ΔTrend (year) | 1.136 | 0.811 | 1.590 |

^1^ Among past-year smokers who made ≥1 quit attempt in the past 12 months.

^a^ Adjusted for seasonality, age, gender, social grade, region, cigarette dependence, and national expenditure on tobacco control mass media campaigns.

### Part 5: References

1	Vangeli E, Stapleton J, Smit ES, Borland R, West R. Predictors of attempts to stop smoking and their success in adult general population samples: a systematic review: Predictors of quit attempts and success. *Addiction* 2011; **106**: 2110–21.

2	Kuipers MAG, Beard E, West R, Brown J. Associations between tobacco control mass media campaign expenditure and smoking prevalence and quitting in England: a time series analysis. *Tob Control* 2018; **27**: 455–62.

3	Beard E, Jackson SE, West R, Kuipers MAG, Brown J. Population-level predictors of changes in success rates of smoking quit attempts in England: a time series analysis. *Addiction* 2020; **115**: 315–25.

4	Langley T, Szatkowski L, Lewis S, *et al.* The freeze on mass media campaigns in England: a natural experiment of the impact of tobacco control campaigns on quitting behaviour. *Addict Abingdon Engl* 2014; **109**: 995–1002.

5	Fidler JA, Shahab L, West R. Strength of urges to smoke as a measure of severity of cigarette dependence: comparison with the Fagerström Test for Nicotine Dependence and its components. *Addict Abingdon Engl* 2011; **106**: 631–8.

6	Montgomery JM, Nyhan B, Torres M. How Conditioning on Posttreatment Variables Can Ruin Your Experiment and What to Do about It. *Am J Polit Sci* 2018; **62**: 760–75.
